# Sequential Organ Failure Assessment (SOFA) score for predicting mortality in patients with sepsis in Vietnamese intensive care units: A multicentre, cross-sectional study

**DOI:** 10.1101/2022.05.18.22275206

**Authors:** Son Ngoc Do, Co Xuan Dao, Tuan Anh Nguyen, My Ha Nguyen, Dung Thi Pham, Nga Thi Nguyen, Dai Quang Huynh, Quoc Trong Ai Hoang, Cuong Van Bui, Thang Dinh Vu, Ha Nhat Bui, Hung Tan Nguyen, Hai Bui Hoang, Thuy Thi Phuong Le, Lien Thi Bao Nguyen, Phuoc Thien Duong, Tuan Dang Nguyen, Vuong Hung Le, Giang Thi Tra Pham, Giang Thi Huong Bui, Tam Van Bui, Thao Thi Ngoc Pham, Chi Van Nguyen, Anh Dat Nguyen, Jason Phua, Andrew Li, Chinh Quoc Luong

## Abstract

**Objectives:** To compare the accuracy of the SOFA and APACHE II scores in predicting mortality among ICU patients with sepsis in an LMIC.

**Design:** A multicentre, cross-sectional study.

**Setting:** A total of 15 adult ICUs throughout Vietnam.

**Participants:** We included all patients aged ≥ 18 years who were admitted to ICUs for sepsis and who were still in ICUs from 00:00 hour to 23:59 hour of the specified study days (i.e., 9^th^ January, 3^rd^ April, 3^rd^ July, and 9^th^ October of the year 2019).

**Primary and secondary outcome measures:** The primary outcome was hospital all-cause mortality (hospital mortality). We also defined the secondary outcome as all-cause deaths in the ICU (ICU mortality).

**Results:** Of 252 patients, 40.1% died in hospitals, and 33.3% died in ICUs. SOFA (AUROC: 0.688 [95% CI: 0.618-0.758]; cut-off value ≥ 7.5; P_AUROC_<0.001) and APACHE II scores (AUROC: 0.689 [95% CI: 0.622-0.756]; cut-off value ≥ 20.5; P_AUROC_<0.001) both had a poor discriminatory ability for predicting hospital mortality. However, the discriminatory ability for predicting ICU mortality of SOFA (AUROC: 0.713 [95% CI: 0.643-0.783]; cut-off value ≥ 9.5; P_AUROC_<0.001) was fair and was better than that of APACHE II score (AUROC: 0.672 [95% CI: 0.603-0.742]; cut-off value ≥ 18.5; P_AUROC_<0.001). A SOFA score ≥ 8 (adjusted OR: 2.717; 95% CI: 1.371-5.382) and an APACHE II score ≥ 21 (adjusted OR: 2.668; 95% CI: 1.338-5.321) were independently associated with an increased risk of hospital mortality. Additionally, a SOFA score ≥ 10 (adjusted OR: 2.194; 95% CI: 1.017-4.735) was an independent predictor of ICU mortality, in contrast to an APACHE II score ≥ 19, for which this role did not.

**Conclusions:** In this study, SOFA and APACHE II scores were worthwhile in predicting mortality among ICU patients with sepsis. However, due to better discrimination for predicting ICU mortality, the SOFA was preferable to the APACHE II score in predicting mortality.

Clinical trials registry – India: CTRI/2019/01/016898

**Strengths and limitations of this study:**

- An advantage of the present study was data from multi centres, which had little missing data.
- Due to the absence of a national registry of intensive care units (ICUs) to allow systematic recruitment of units, we used a snowball method to identify suitable units, which might have led to the selection of centres with a greater interest in sepsis management.
- Due to the study’s real-world nature, we did not make a protocol for microbiological investigations. Moreover, we mainly evaluated resources utilized in ICUs; therefore, the data detailing the point-of-care testing and life-sustaining treatments were not available. Additionally, to improve the feasibility of conducting the study in busy ICUs, we opted not to collect data on antibiotic resistance and appropriateness.
- Due to our independent variables (e.g., SOFA score that was greater than or equal to the cut-off value) that might be associated with primary outcome only measured upon ICU admission, the mixed-effects logistic regression model could not be used to predict discrete outcome variables measured at two different times, i.e., inside and outside the ICU settings.
- Although the sample size was large enough, the confidence interval was slightly wide (±6.03%), which might influence the normal distribution of the sample.

## INTRODUCTION

Sepsis is a clinical syndrome which has physiologic, biologic, and biochemical abnormalities caused by a dysregulated host response to infection and is a critical global health problem.(1, 2) Sepsis is the most common cause of in-hospital deaths, with most of the burden in low- and middle-income countries (LMICs), and extracts a high economic and social cost;(3-5) mortality rates remain high at 30-45% and contribute to as much as 20% of all deaths worldwide.(2, 4, 6, 7) There is no reference standard that allows easy, accurate diagnosis and prognosis of sepsis.(1, 8) Although the 1991 International Consensus Definition Task Force proposed the systemic inflammatory response syndrome (SIRS) criteria to identify patients with a septic host response,(9) these criteria do not measure whether the response is injurious, and their utility is limited.(1, 8).

The Acute Physiology and Chronic Health Examination II (APACHE II) score was originally developed for critically ill patients in intensive care units (ICUs).(10) It has 12 physiologic measures and extra points based upon age and the presence of chronic disease.(10) The APACHE II score was shown to have good prognostic value in acutely ill or surgical patients. (10, 11) However, some limitations of the APACHE II score are that (i) it is complex and cumbersome to use, (ii) it does not differentiate between the sterile and infected necrosis, and finally, (iii) it has a poor predictive value at 24 hours.(12)

In 2016, the Sepsis-3 Task Force proposed that for patients with suspected infection, an increase of 2 points or more in the Sequential (Sepsis-Related) Organ Failure Assessment (SOFA) score could serve as clinical criteria for sepsis,(1) and the consensus has not changed since then.(13) This approach was justified based on content validity (SOFA reflects the facets of organ dysfunction) and predictive validity (the proposed criteria predict downstream events associated with the condition of interest).(14-17) However, the validity of this score was mainly derived from critically-ill patients with suspected sepsis by interrogating over a million intensive care unit (ICU) electronic health record encounters from ICUs in high-income countries (HICs).(1, 17, 18) Moreover, the patients, pathogens, and clinical capacity to manage sepsis differ considerably between HIC and LMIC settings.(7) Therefore, it’s still unclear whether this score could be applied to different types of infection, locations within the hospital, and countries.

Vietnam is an LMIC, ranked 15^th^ in the world and 3^rd^ in Southeast Asia by population with 96.462 million people.(19) Vietnam is also a hotspot for emerging infectious diseases in Southeast Asia, including the SARS-CoV,(20) avian influenza A(H5N1),(21, 22) and ongoing global COVID-19 outbreaks(23, 24). Additionally, severe dengue,(25) *Streptococcus suis* infection,(26) malaria,(27) and increased antibiotic resistance are other major causes of sepsis in ICUs across Vietnam(28, 29). Despite its recent economic growth spurt,(30) Vietnam is still struggling to provide either enough resources or adequate diagnostic, prognostic and treatment strategies for patients with sepsis in both local and central settings.(31, 32) In addition, within the healthcare system in Vietnam, central hospitals are responsible for receiving patients who have difficulties being treated in local hospital settings.(33) Therefore, the diagnosis, prognosis, and initiation of treatment for patients with sepsis are often delayed.

In resource-limited settings, the early identification of infected patients who may go on to develop sepsis or may be at risk of death from sepsis using accurate scoring systems as a way to decrease sepsis-associated mortality. Therefore, this study aimed to investigate the mortality rate and compare the accuracy of the SOFA score and the APACHE II score in predicting mortality in ICU patients with sepsis in Vietnam.

## METHODS

### Source of data

This multicentre observational, cross-sectional, point prevalence study is part of the Management of Severe sepsis in Asia’s Intensive Care unitS (MOSAICS) II study,(34-37) which enrolled patients on 9^th^ January (Winter), 3^rd^ April (Spring), 3^rd^ July (Summer), and 9^th^ October (Autumn) of the year 2019. All patients received a follow-up till hospital discharge, death in the ICU/hospital, or up to 90 days post-enrolment, whichever was earliest. In this study, we used only data from Vietnam. A total of 15 adult ICUs (excluding predominantly neurosurgical, coronary, and cardiothoracic ICUs) participating in the MOSAICS II study from 14 hospitals, of which 5 are central and 9 are provincial, district, or private hospitals, throughout Vietnam. Each ICU had one or two representatives who were part of the local study team and the MOSAICS II study group, as shown in eAppendix 2 of a previously published paper(36). Participation was voluntary and unfunded.

### Participants

All patients admitted to participating ICUs on one of the four days (i.e., January 9^th^, April 3^rd^, July 3^rd^, and October 9^th^, 2019) which represented the different seasons of the year 2019 were screened for eligibility. We included all patients, aged ≥ 18 years old, who were admitted to the ICUs for sepsis, and who were still in the ICUs from 00:00 hour to 23:59 hour of the study days. We defined sepsis as infection with a SOFA score of 2 points or more from baseline (assumed to be 0 for patients without prior organ dysfunction).(1)

### Data collection

We used a standardized classification and case record form (CRF) to collect data on common variables as shown in Supplementary file 1. The data dictionary of the MOSAICS II study is available as an online supplement of previously published papers.(35, 36) Data was entered by the representatives of the participating hospitals into the database of the MOSAICS II study via the password-protected online CRFs. We checked the data for implausible outliers and missing fields and contacted ICU representatives for clarification. We then merged the data sets for the 14 hospitals.

### Outcome measures

The primary outcome was hospital all-cause mortality (hospital mortality). We also defined the secondary outcomes as all-cause deaths in the ICU (ICU mortality) and the ICU and hospital lengths of stay (LOS).

### Predictor measures

We defined exposure variables as the SOFA and the APACHE II scores.(10, 14) All data elements required for calculating the SOFA score at the time of ICU admission and the APACHE II score over the first 24 hours of ICU admission were prospectively collected on a CRF and entered into a database via the online CRF for later analysis.

We determined confounding factors as the variables of hospital and ICU characteristics collected on a questionnaire by representatives before patient enrolment, as shown in Supplementary file 2. We also determined confounding factors as variables collected on a CRF by investigators. The CRF contained 4 sections which is available in Supplementary file 1. The first section focused on baseline characteristics (demographics, documented comorbidities, and details of admission). The second section comprised of vital signs upon ICU admission, laboratory parameters, site of infection, and microbiology. Only microorganisms detected via all cultures, serology, molecular, and histological investigations and deemed to be true pathogens rather than commensals or contaminants were recorded. The third section captured the timing of sepsis bundle elements referencing time zero, determined as follows: (a) time of triage in the emergency department (ED) for those presenting with sepsis to the ED; (b) time of clinical documentation of deterioration in the general wards or other non-ED areas for those who developed sepsis after hospital admission; (c) time of ICU admission for those in which (a) or (b) could not be determined from the clinical documentation. The bundle elements were based on the Surviving Sepsis Campaign’s 2018 update: antibiotics administration, blood cultures, lactate measurement, fluid administration (amount of fluids administered in the first and third hours from time zero) and vasopressor initiation.(38) The fourth section concerned life-sustaining treatments provided during the ICU stay.

### Sample size

In the present study, hospital mortality served as the primary outcome. We, therefore, used the formula to determine the minimal sample size for estimating a population proportion with a confidence level of 95%, a confidence interval (margin of error) of 6.03% and an assumed population proportion of 61.0%, based on the hospital mortality rate (61.0%) of our cohort reported in a previously published study(39). Therefore, we should have at least 252 patients in our sample. Because of this, our sample size was sufficient and reflected a normal distribution.

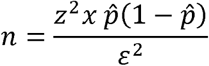

where:

*z is the z score* (*z score for a* 95% *confidence level is* 1.96)

*E is the margin of error* (ε *for a confidence interval of ±* 6.03% *is* 0.0603)

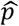 *is the population proportion* (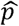. *for a population proportion of* 61.0% *is* 0.61) *n is the sample size*

### Statistical analyses

We used IBM^®^ SPSS^®^ Statistics 22.0 (IBM Corp., Armonk, United States of America) for data analysis. We report data as numbers (no.) and percentages (%) for categorical variables and medians and interquartile ranges (IQRs) or means and standard deviations (SDs) for continuous variables. Comparisons were made between survival and death in the hospital and ICU for each variable, using the Chi-squared test or Fisher exact test for categorical variables and the Mann-Whitney U test, Kruskal-Wallis test, one-way analysis of variance for continuous variables.

Receiver operator characteristic (ROC) curves were plotted and the areas under the receiver operating characteristic curve (AUROC) were calculated to determine the discriminatory ability of the SOFA and APACHE II scores for deaths in the hospital and ICU. The cut-off value of the SOFA and the APACHE II scores was determined by the ROC curve analysis and defined as the cut-off point with the maximum value of Youden’s index (i.e., sensitivity + specificity – 1). Based on the cut-off value of the scores, we assigned the patients to two groups: either a score that was less than the cut-off value or a score that was greater than or equal to the cut-off value.

We assessed factors associated with death in the hospital using logistic regression analysis. To reduce the number of predictors and the multicollinearity issue and resolve the overfitting, we used different ways to select variables as follows: (a) we put all variables (including exposure and confounding factors) of hospital and ICU characteristics, baseline characteristics, clinical and laboratory characteristics, and treatments into the univariable logistic regression model; (b) we selected variables if the P-value was <0.05 in the univariable logistic regression analysis between survival and death in the hospital, as well as those that are clinically crucial to put in the multivariable logistic regression model. These variables included university affiliation, training program in ICU, documented comorbidities (i.e., cardiovascular disease, chronic neurological disease), the severity of illness (i.e., SOFA and APACHE II scores that were greater than or equal to the cut-off value), sites of infection (i.e., urinary tract, abdominal, skin or cutaneous sites), pathogens detection (i.e., no pathogens detected, Gram-negative bacteria), completion of the 1- or 3-hour sepsis bundle of care, completion of the initial administration of antibiotics within 1 or 3 hours, respiratory support (i.e., mechanical ventilation (MV), high-flow nasal oxygen), and additional ICU support (i.e., vasopressors/inotropes, renal replacement therapy (RRT), red blood cell transfusion, platelet transfusion, fresh frozen plasma transfusion, surgical source control, and non-surgical source control). Using a stepwise backward elimination method, we started with the full multivariable logistic regression model that included the selected variables. This method then deleted the variables stepwise from the full model until all remaining variables were independently associated with the risk of death in the hospital in the final model. Similarly, we used these methods of variable selection and analysis for assessing factors associated with death in the ICU. We presented the odds ratios (ORs) and 95% confidence intervals (CIs) in the univariable logistic regression model and the adjusted odds ratios (AORs) and 95% CIs in the multivariable logistic regression model.

For all analyses, significance levels were two-tailed, and we considered p□<0.05 as statistically significant.

### Patient and public involvement

Patients or the public were not involved in the design, or conduct, or reporting, or dissemination plans of our research.

## RESULTS

Data on 252 patients with sepsis were submitted to the database of the MOSAICS II study (Fig. 1), in which there were little missing data.

**Figure 1.**
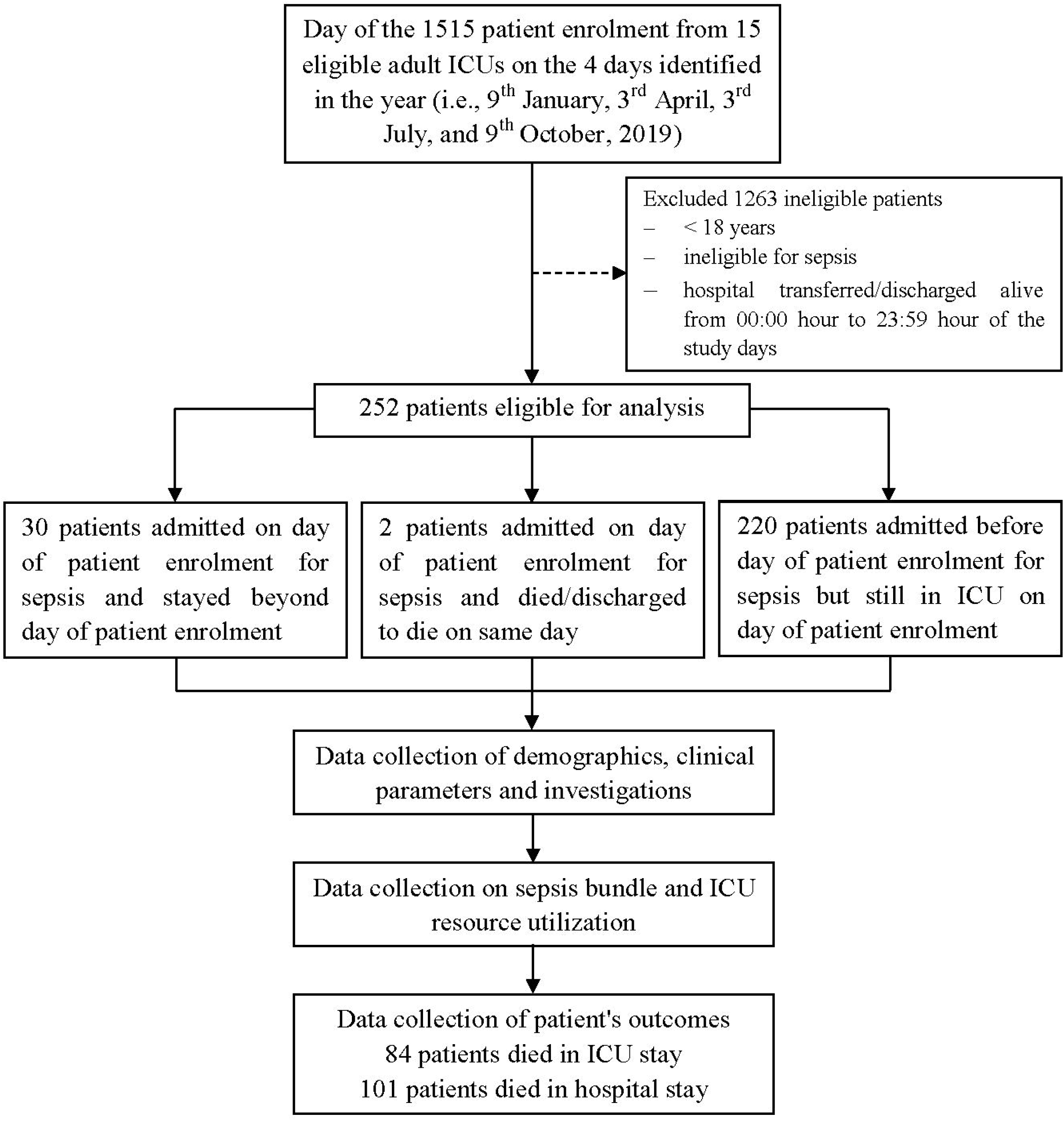
Flowchart of the study design, patient enrolment and follow-up (<u>Abbreviations</u>: **ICU**, intensive care unit; **“discharged to die”**, defined as the patients were in grave condition or dying and were classified with deaths in the ICU at the time of discharge).

### Clinical characteristics and outcomes

In our study cohort, 64.3% (162/252) were men and the median age was 65 years (IQR: 52– 76.75) (Table 1). Among the total patients, the median SOFA score was 7 (IQR: 4.75–10) at the time of ICU admission, the median APACHE II score was 18 (IQR: 13– 24) over the first 24 hours of ICU admission, and 29.4% (74/252) of patients had septic shock (Table 1). Table 1 also shows that the most common documented comorbidities included cardiovascular disease (31.0%; 78/252), diabetes mellitus (26.6%; 67/252), and chronic neurological disease (14.3%; 36/252), the most common sites of infection included respiratory (56.7%; 143/252), abdominal cavity (24.2%; 61/252), urinary tract (14.7%; 37/252) and skin or cutaneous sites (7.5%; 19/252) and Gram-negative bacteria were isolated in 61.9% (156/252) of patients. Table 2 shows that MV was provided for 68.9% (173/251) of patients and RRT for 40.2% (101/251). Overall, 40.1% (101/252) of patients with sepsis died in the hospital, 33.3% (84/252) of whom died in the ICU (Fig. 1 and Table 2). The median hospital and ICU LOS were 16 (IQR: 10–25) and 10 (IQR: 6–18) days, respectively (Table 2). The clinical characteristics, severity of illness, sites of infection and microbiology, compliance with sepsis bundle elements, and life-sustaining treatments during ICU stay were compared between patients who survived and patients who died in the hospital and ICU, as shown in Tables 1 and 2, and Tables S1–S14 (Supplementary file 3).

**Table 1.** Baseline characteristics according to hospital survivability of patients with sepsis

| Variables | All cases | Survived | Died | p<sup>a</sup> |
| --- | --- | --- | --- | --- |
| **Hospital and ICU characteristics** | n=252 | n=151 | n=101 |  |
| University affiliation, no. (%) | 99 (39.3) | 46 (30.5) | 53 (52.5) | <0.001 |
| Training programme in ICU, no. (%) | 202 (80.2) | 129 (85.4) | 73 (72.3) | 0.010 |
| **Demographics** | n=252 | n=151 | n=101 |  |
| Age (year), median (IQR) | 65 (52-76.75) | 65 (53-76) | 65 (52-78) | 0.810<sup>**</sup> |
| Sex (male), no. (%) | 162 (64.3) | 93 (61.6) | 69 (68.3) | 0.275 |
| **Documented comorbidities** | n=252 | n=151 | n=101 |  |
| Cardiovascular disease, no. (%) | 78 (31.0) | 41 (27.2) | 37 (36.6) | 0.111 |
| Chronic lung disease, no. (%) | 30 (11.9) | 18 (11.9) | 12 (1.9) | 0.992 |
| Chronic neurological disease, no. (%) | 36 (14.3) | 28 (18.5) | 8 (7.9) | 0.018 |
| Chronic kidney disease, no. (%) | 23 (9.1) | 14 (9.3) | 9 (8.9) | 0.922 |
| Peptic ulcer disease, no. (%) | 9 (3.6) | 5 (3.3) | 4 (4.0) | >0.999<sup>*</sup> |
| Chronic liver disease, no. (%) | 27 (10.7) | 14 (9.3) | 13 (12.9) | 0.365 |
| Diabetes mellitus, no. (%) | 67 (26.6) | 40 (26.5) | 27 (26.7) | 0.966 |
| Connective tissue disease, no. (%) | 3 (1.2) | 2 (1.3) | 1 (1.0) | >0.999<sup>*</sup> |
| Immunosuppression, no. (%) | 10 (4.0) | 7 (4.6) | 3 (3.0) | 0.744 |
| Haematological malignancies, no. (%) | 5 (2.0) | 3 (2.0) | 2 (2.0) | >0.999<sup>*</sup> |
| Solid malignant tumours, no. (%) | 12 (4.8) | 6 (4.0) | 6 (5.9) | 0.551<sup>*</sup> |
| **Vital signs (on admission into ICU)** | n=252 | n=151 | n=101 |  |
| GCS, median (IQR) | 13 (9-15) | 14 (10-15) | 10 (8-14) | <0.001<sup>**</sup> |
| HR (beats per min), median (IQR) | 110 (95.25-125.75) | 110 (92-125) | 110 (100-129.5) | 0.083<sup>**</sup> |
| Temperature (°C), mean (SD) | 37.79 (1.01) | 37.80 (1.08) | 37.77 (0.91) | 0.871<sup>**</sup> |
| MBP (mmHg), mean(SD) | 75.82 (22.08) | 79.75 (22.88) | 69.93 (19.51) | 0.002<sup>**</sup> |
| RR (breaths per min), median (IQR) | 25 (22-30) | 25 (22-30) | 25 (20-30) | 0.693<sup>**</sup> |
| **Blood investigations** | n=252 | n=151 | n=101 |  |
| Total WBC (x10<sup>9</sup>/L), mean (SD) | 15.73 (9.20) | 15.63 (8.67) | 15.88 (9.98) | 0.914<sup>**</sup> |
| PLT (x10<sup>9</sup>/L), mean (SD) | 185.98 (137.85) | 200.71 (129.67) | 163.95 (147.15) | 0.002<sup>**</sup> |
| Hb (g/dL), mean (SD) | 11.14 (2.59) | 11.36 (2.68) | 10.82 (2.44) | 0.088<sup>**</sup> |
| K<sup>+</sup> (mmol/L), mean (SD) | 3.89 (0.79) | 3.90 (0.80) | 3.87 (0.77) | 0.865<sup>**</sup> |
| Na<sup>+</sup> (mmol/L), mean (SD) | 136.05 (8.24) | 135.62 (8.81) | 136.69 (7.80) | 0.068<sup>**</sup> |
| Creatinine (µmol/L), mean (SD) | 187.85 (151.92) | 186.15 (171.60) | 190.38 (117.27) | 0.030<sup>**</sup> |
| Bilirubin (µmol/l), mean (SD) | 32.80 (61.49) | 31.74 (72.67) | 34.35 (40.09) | 0.007<sup>**</sup> |
| pH, mean (SD) | 7.37 (0.50) | 7.41 (0.64) | 7.32 (0.14) | 0.004<sup>**</sup> |
| PaO<sub>2</sub> (mmHg), mean (SD) | 116.17 (74.28) | 110.23 (56.25) | 124.73 (94.07) | 0.665<sup>**</sup> |
| PaO<sub>2</sub>/FiO<sub>2</sub> ratio, mean (SD) | 262.48 (149.58) | 281.52 (149.39) | 235.26 (146.32) | 0.003<sup>**</sup> |
| **Severity of illness scores** | n=252 | n=151 | n=101 |  |
| SOFA, median (IQR), n=250 | 7 (4.75-10) | 6 (4-9) | 9 (6-12) | <0.001<sup>**</sup> |
| APACHE II, median (IQR) | 18 (13-24) | 15 (12-21) | 22 (16-27) | <0.001<sup>**</sup> |
| Septic Shock | 74 (29.4) | 35 (23.2) | 39 (38.6) | 0.008 |
| **Site of Infection** | n=252 | n=151 | n=101 |  |
| Respiratory, no. (%) | 143 (56.7) | 82 (54.3) | 61 (60.4) | 0.339 |
| Urinary tract, no. (%) | 37 (14.7) | 30 (19.9) | 7 (6.9) | 0.004 |
| Abdominal, no. (%) | 61 (24.2) | 34 (22.5) | 27 (26.7) | 0.444 |
| Neurological, no. (%) | 12 (4.8) | 8 (5.3) | 4 (4.0) | 0.767* |
| Bones or joints, no. (%) | 2 (0.8) | 2 (1.3) | 0 (0.0) | 0.518* |
| Skin or cutaneous sites, no. (%) | 19 (7.5) | 7 (4.6) | 12 (11.9) | 0.033 |
| Intravascular catheter, no. (%) | 1 (0.4) | 1 (0.7) | 0 (0.0) | >0.999* |
| Infective endocarditis, no. (%) | 1 (0.4) | 0 (0.0) | 1 (1.0) | 0.401* |
| Primary bacteraemia, no. (%) | 7 (2.8) | 5 (3.3) | 2 (2.0) | 0.705* |
| Systemic, no. (%) | 6 (2.4) | 4 (2.6) | 2 (2.0) | >0.999* |
| **Microbiology** | n=252 | n=151 | n=101 |  |
| No pathogens detected, no. (%) | 67 (26.6) | 47 (31.1) | 20 (19.8) | 0.046 |
| Gram negative bacteria, no. (%) | 156 (61.9) | 88 (58.3) | 68 (67.3) | 0.147 |
| Gram positive bacteria, no. (%) | 34 (13.5) | 22 (14.6) | 12 (11.9) | 0.540 |
| Fungi, no. (%) | 7 (2.8) | 4 (2.6) | 3 (3.0) | >0.999* |
| Viruses, no. (%) | 2 (0.8) | 0 (0.0) | 2 (2.0) | 0.160* |
| Other pathogens, no. (%) | 4 (1.6) | 3 (2.0) | 1 (1.0) | 0.651* |

\*Comparison between the patients who survived and died using Chi-squared test; \*Fisher's exact test; \*\*Mann-Whitney U test.

**Table 2.**
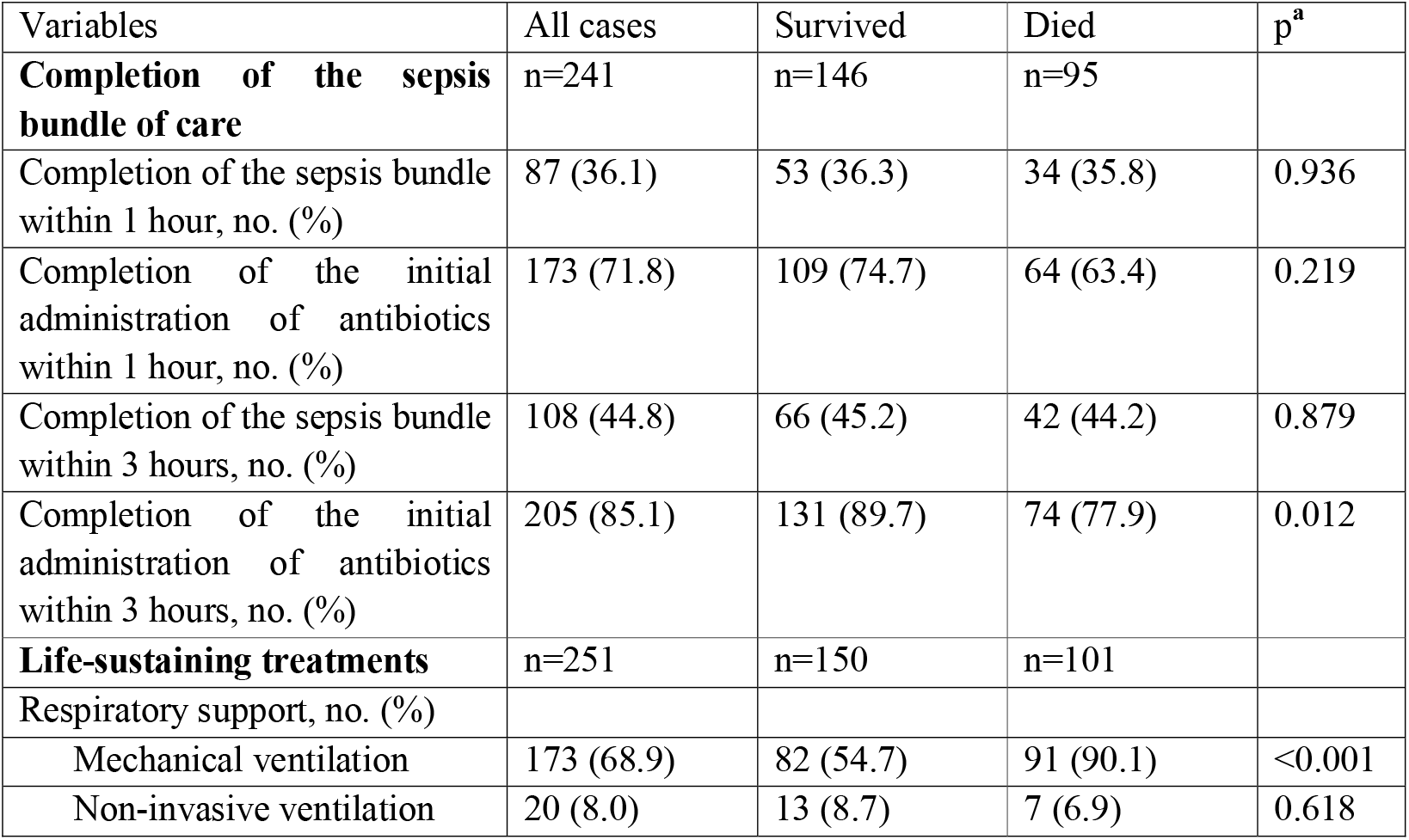
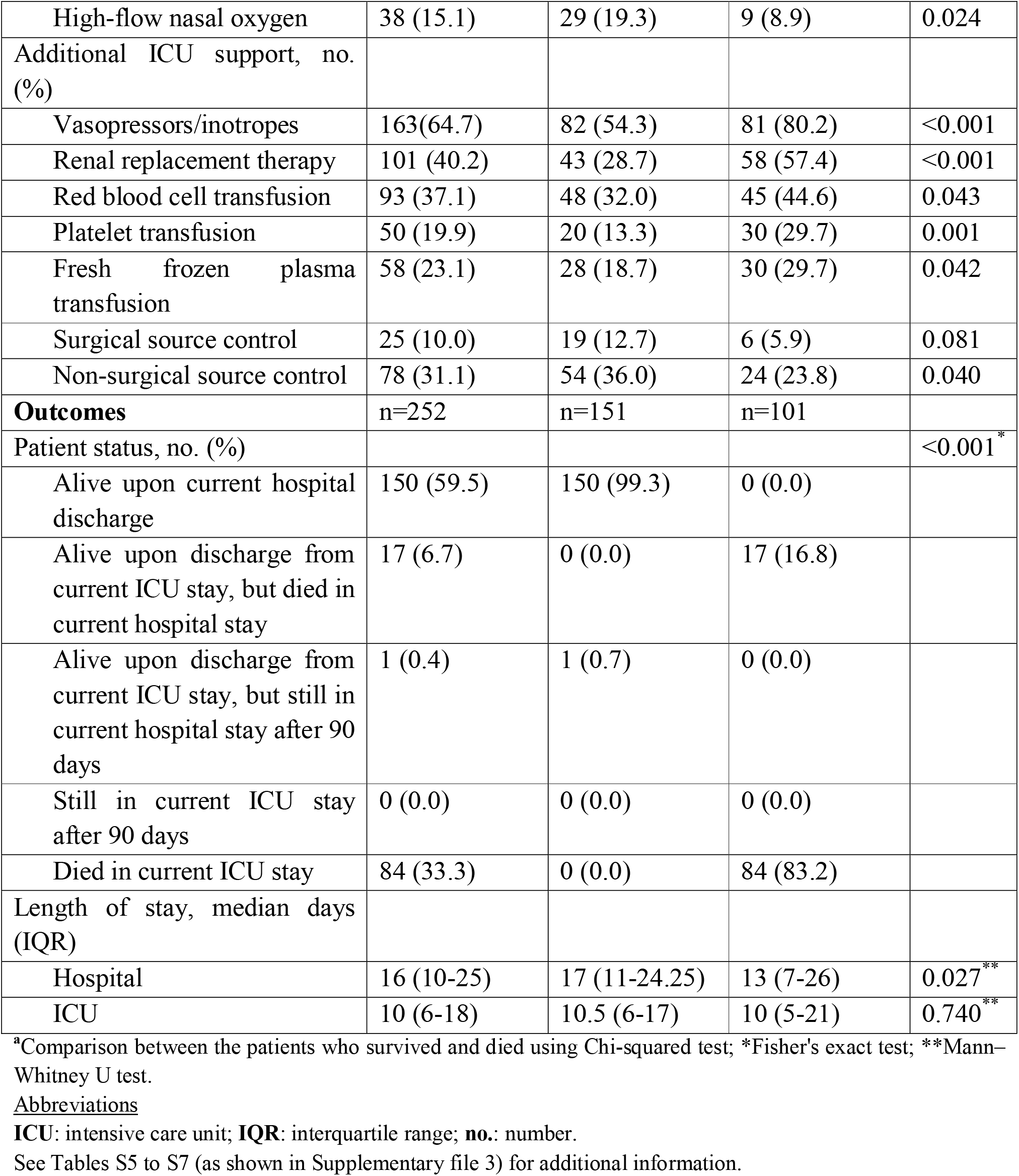
Treatments and outcomes according to hospital survivability of patients with sepsis

### Overall prognostic performance of the severity scoring systems

The SOFA score (AUROC: 0.688 [95% CI: 0.618–0.758]; cut-off value ≥ 7.5; sensitivity: 64.4%; specificity: 69.8%; P_AUROC_ <0.001) and APACHE II score (AUROC: 0.689 [95% CI: 0.622–0.756]; cut-off value ≥ 20.5; sensitivity: 61.4%; specificity: 71.8%; P_AUROC_ <0.001) both had a poor discriminatory ability for the hospital mortality (Fig. 2). The discriminatory ability for the ICU mortality of SOFA score (AUROC: 0.713 [95% CI: 0.643–0.783]; cut-off value ≥ 9.5; sensitivity: 53.6%; specificity: 80.1%; P_AUROC_ <0.001), however, was fair and was better than that of the APACHE II score (AUROC: 0.672 [95% CI: 0.603–0.742]; cut-off value ≥ 18.5; sensitivity: 69.0%; specificity: 60.8%; P_AUROC_ <0.001) (Fig. 3).

**Figure 2.**
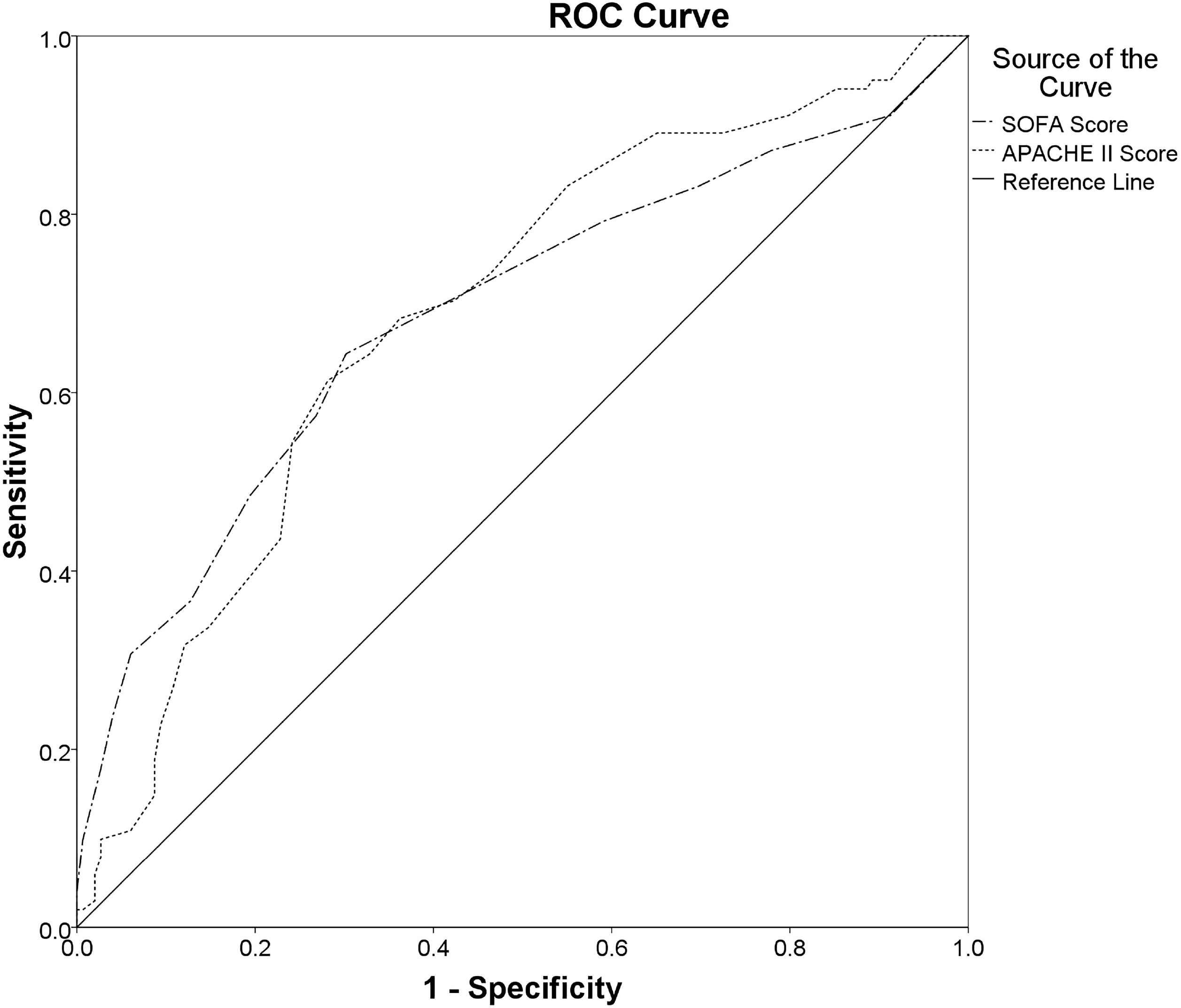
Comparisons of the AUROCs: Comparing the overall diagnostic performance of the SOFA score (AUROC: 0.688 [95% CI: 0.618–0.758]; cut-off value ≥ 7.5; sensitivity: 64.4%; specificity: 69.8%; P_AUROC_ <0.001) and the APACHE II score (AUROC: 0.689 [95% CI: 0.622–0.756]; cut-off value ≥ 20.5; sensitivity: 61.4%; specificity: 71.8%; P_AUROC_ <0.001) for predicting hospital mortality in ICU patients with sepsis (<u>Abbreviations</u>: **APACHE II**: Acute Physiology and Chronic Health Evaluation II score; **AUROC**: areas under the receiver operating characteristic curve; **CI**: confidence interval; **ICU**: intensive care unit; **ROC**: receiver operator characteristic curve; **SOFA**: Sequential Organ Failure Assessment score).

**Figure 3.**
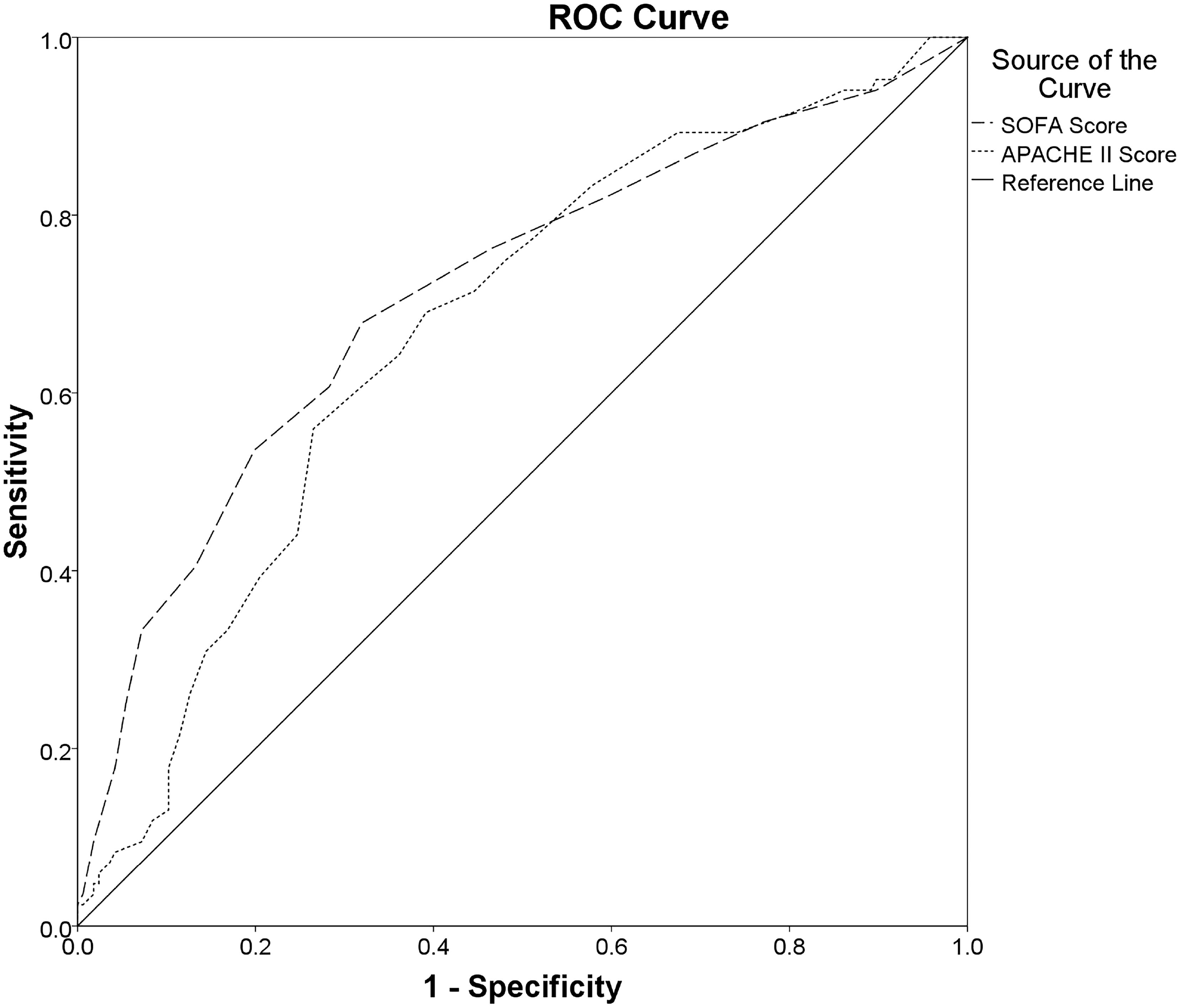
Comparisons of the AUROCs: Comparing the overall diagnostic performance of the SOFA score (AUROC: 0.713 [95% CI: 0.643–0.783]; cut-off value ≥ 9.5; sensitivity: 53.6%; specificity: 80.1%; P_AUROC_ <0.001) and the APACHE II score (AUROC: 0.672 [95% CI: 0.603–0.742]; cut-off value ≥ 18.5; sensitivity: 69.0%; specificity: 60.8%; P_AUROC_ <0.001) for predicting ICU mortality in ICU patients with sepsis (<u>Abbreviations</u>: **APACHE II**: Acute Physiology and Chronic Health Evaluation II score; **AUROC**: areas under the receiver operating characteristic curve; **CI**: confidence interval; **ICU**: intensive care unit; **ROC**: receiver operator characteristic curve; **SOFA**: Sequential Organ Failure Assessment score).

### Risk factors for mortality

In the multivariable analysis, a SOFA score of 8 and above (AOR: 2.717; 95% CI: 1.371– 5.382) and an APACHE II score of 21 and above (AOR: 2.668; 95% CI: 1.338–5.321) that were independently associated with an increased risk of hospital mortality (Table 3). Additionally, a SOFA score of 10 and above (AOR: 2.801; 95% CI: 1.332–5.891) was independently associated with an increased risk of ICU mortality, in contrast to an APACHE II score of 19 and above, for which this independent association was not observed (Table 4). Other factors were significantly or independently associated with the risk of hospital and ICU mortalities, as shown in Tables 3 and 4, and Tables S15–S18 (Supplementary file 3).

**Table 3.** Factors relating to hospital mortality in patients with sepsis

| Factors | Univariable logistic regression analyses<sup>a</sup> |  |  |  | Multivariable logistic regression analyses<sup>b</sup> |  |  |  |
| --- | --- | --- | --- | --- | --- | --- | --- | --- |
|  | OR | 95% CI for OR |  | p value | AOR | 95% CI for AOR |  | p value |
|  |  | Lower | Upper |  |  | Lower | Upper |  |
| **Hospital and ICU characteristics** |  |  |  |  |  |  |  |  |
| University affiliation | 2.520 | 1.495 | 4.248 | 0.001 | NA | NA | NA | NA |
| Training program in ICU | 0.445 | 0.237 | 0.833 | 0.011 | 0.392 | 0.162 | 0.949 | 0.038 |
| **Documented comorbidities** |  |  |  |  |  |  |  |  |
| Cardiovascular disease | 1.551 | 0.903 | 2.664 | 0.112 | 2.181 | 1.019 | 4.664 | 0.044 |
| Chronic neurological disease | 0.378 | 0.165 | 0.867 | 0.022 | 0.179 | 0.058 | 0.546 | 0.003 |
| **Severity of illness scores** |  |  |  |  |  |  |  |  |
| SOFA score ≥ 8 | 4.173 | 2.440 | 7.137 | <0.001 | 2.717 | 1.371 | 5.382 | 0.004 |
| APACHE II score ≥ 21 | 4.126 | 2.414 | 7.051 | <0.001 | 2.668 | 1.338 | 5.321 | 0.005 |
| **Site of infection** |  |  |  |  |  |  |  |  |
| Urinary tract | 0.300 | 0.126 | 0.714 | 0.006 | 0.312 | 0.105 | 0.932 | 0.037 |
| Abdominal | 1.256 | 0.701 | 2.249 | 0.444 | NA | NA | NA | NA |
| Skin or cutaneous sites | 2.774 | 1.053 | 7.309 | 0.039 | NA | NA | NA | NA |
| **Microbiology** |  |  |  |  |  |  |  |  |
| No pathogens detected | 0.546 | 0.300 | 0.994 | 0.048 | NA | NA | NA | NA |
| Gram-negative bacteria | 1.475 | 0.871 | 2.498 | 0.148 | NA | NA | NA | NA |
| **Completion of sepsis bundle elements** |  |  |  |  |  |  |  |  |
| Completion of the sepsis bundle within 1 hour | 0.978 | 0.571 | 1.675 | 0.936 | NA | NA | NA | NA |
| Completion of the administration of antibiotics within 1 hour | 0.701 | 0.397 | 1.237 | 0.220 | NA | NA | NA | NA |
| Completion of the sepsis bundle within 3 hours | 0.961 | 0.571 | 1.615 | 0.879 | NA | NA | NA | NA |
| Completion of the administration of antibiotics within 3 hours | 0.403 | 0.196 | 0.830 | 0.014 | 0.381 | 0.151 | 0.965 | 0.042 |

| **Life-sustaining treatments during ICU stay** |  |  |  |  |  |  |  |  |
| --- | --- | --- | --- | --- | --- | --- | --- | --- |
| *Respiratory support* |  |  |  |  |  |  |  |  |
| Mechanical ventilation | 7.546 | 3.645 | 15.625 | <0.001 | 4.391 | 1.912 | 10.085 | <0.001 |
| High-flow nasal oxygen | 0.408 | 0.184 | 0.904 | 0.027 | NA | NA | NA | NA |
| *Additional ICU support* |  |  |  |  |  |  |  |  |
| Vasopressors/inotropes | 3.408 | 1.899 | 6.116 | <0.001 | NA | NA | NA | NA |
| Renal replacement therapy | 3.356 | 1.976 | 5.702 | <0.001 | NA | NA | NA | NA |
| Red blood cell transfusion | 1.708 | 1.014 | 2.876 | 0.044 | NA | NA | NA | NA |
| Platelet transfusion | 2.746 | 1.455 | 5.185 | 0.002 | NA | NA | NA | NA |
| Fresh frozen plasma transfusion | 1.841 | 1.018 | 3.329 | 0.043 | NA | NA | NA | NA |
| Surgical source control | 0.435 | 0.168 | 1.132 | 0.088 | NA | NA | NA | NA |
| Non-surgical source control | 0.554 | 0.314 | 0.977 | 0.041 | NA | NA | NA | NA |
| Constant |  |  |  |  | 0.230 |  |  | 0.007 |

<sup>a</sup>Each variable of hospital and ICU characteristics, baseline characteristics, clinical and laboratory characteristics, and treatments was analysed in the univariable logistic regression model and was considered in the multivariable logistic regression model if the P-value was <0.05 in univariable logistic regression analysis between survival and death in the hospital, as well as clinically crucial factors.
<sup>b</sup>All selected variables were included in the multivariable logistic regression model with the stepwise backward elimination method. Variables, then, were deleted stepwise from the full model until all remaining variables were independently associated with death in the hospital.
Abbreviations: **AOR:** adjusted odds ratio; **APACHE II**, Acute Physiology and Chronic Health Examination II; **CI:** confidence interval; **ICU**, intensive care unit; **NA:** not available; **OR:** odds ratio; **SOFA**, Sequential (Sepsis-Related) Organ Failure Assessment.
See Tables S15 and S16 (as shown in Supplementary file 3) for additional information.

**Table 4.**
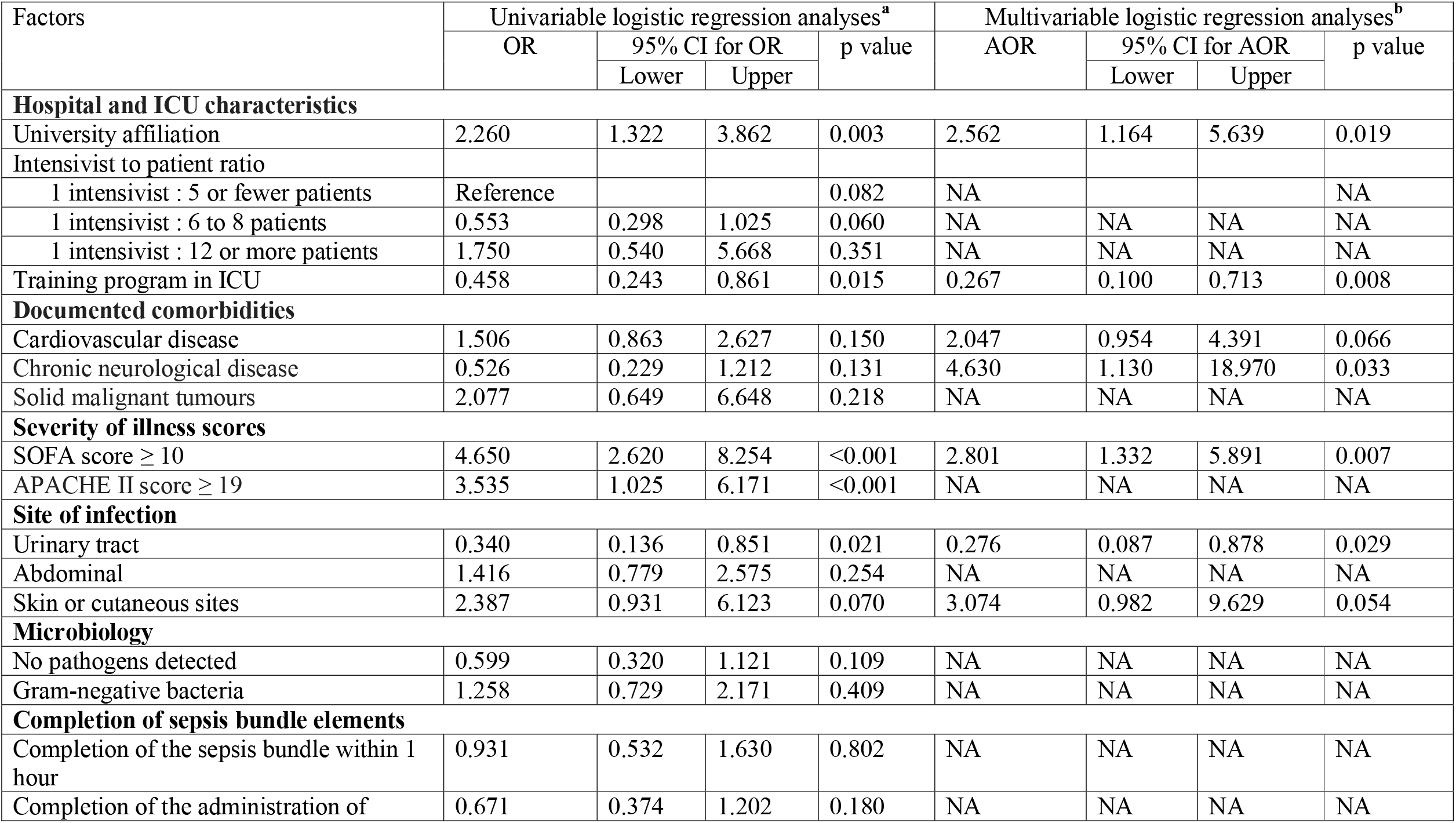
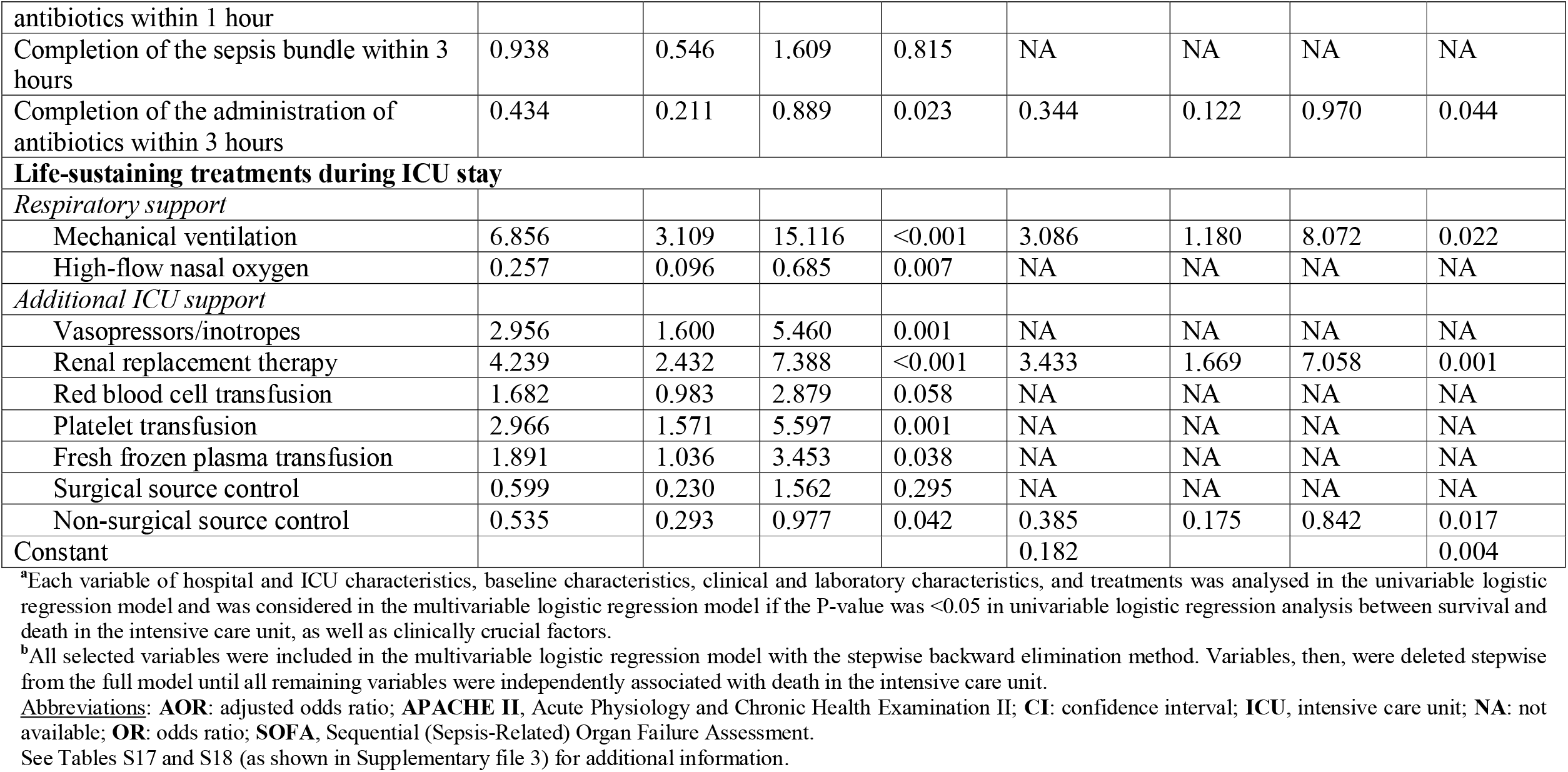
Factors relating to intensive care unit mortality in patients with sepsis

## DISCUSSION

Of 252 patients with sepsis included in our analysis, two-fifths (40.1%) died in the hospital, and about a third (33.3%) died during the ICU stay (Fig. 1 and Table 2). The SOFA and APACHE II scores had a poor discriminatory ability for predicting hospital mortality (Fig. 2). However, the overall performance of the SOFA score for predicting ICU mortality was fair and was better than that of the APACHE II score (Fig. 3). A SOFA score of 8 and above and an APACHE II score of 21 and above were independently associated with an increased risk of hospital mortality (Table 3). Additionally, a SOFA score of 10 and above was an independent predictor of ICU mortality, in contrast to an APACHE II score of 19 and above, for which this role did not (Table 4).

In our study, the hospital mortality rate was lower than that of the MOSAICS I study (44.5%; 572/1285),(40) as well as the rates previously reported from LMICs in Southeast Asia, including Indonesia (68.3%; 41/60),(41) Thailand (42%; 263/627),(42) and Vietnam (61.0%; 75/123)(39). These findings may be because the diagnosis and treatment of sepsis have significantly changed over the previous ten years to increase patient survival in sepsis and septic shock.(1, 8, 13, 36, 38, 43, 44) However, our study showed rates for ICU and hospital mortality that were higher than rates reported in the international Extended Study on Prevalence of Infection in Intensive Care (EPIC III) study (28% [99/352] and 31.1% [110/352] in LMICs, 26.4% [821/3114] and 32.7% [1019/3114] in upper-middle-income countries [UMICs], and 21.3% [950/4470] and 28.5% [1275/4470] in HICs)(45). These variations might be because the EPIC III study included ICU-acquired infection rather than only sepsis.(45) Despite the distinct inclusion criteria, our median SOFA score at the time of ICU admission was comparable to that of the EPIC III study (7 points [IQR: 4–11] in LMICs/UMICs/HICs)(45). However, patients in our study received invasive organ support treatments (i.e., MV and RRT) during ICU stays more frequently than those in the EPIC III study (54.4% [4377/8045] and 15.7% [1253/8045])(45). Previous studies showed that MV was a crucial predictor of mortality at any point throughout the ICU stay.(4, 35) Additionally, the utilization of RRT at any time during the ICU stay was also associated with a higher fatality rate.(4, 35, 46-48) Furthermore, *Acinetobacter baumannii* (17.9%, 45/252; Table S4 as shown in Supplementary file 3), one of the most harmful pathogens, was more frequently isolated from patients in the present study than those from the HIC cohort (4.4%; 137/3113) of the EPIC III study (45). The previous studies showed that *Acinetobacter baumannii* infection was often due to a lack of strict infection control bundles(49) and associated with an increased risk of death(50, 51). The fact that our proportions for ICU and hospital mortality were higher than those reported in the EPIC III study suggested that patients, pathogens, and clinical capacity to manage sepsis vary significantly between regions, particularly between HIC and LMIC settings.

In this study, we found a poor ability of both SOFA and APACHE II scores to predict hospital mortality (Fig. 2). However, with the SOFA score, the discrimination for predicting ICU mortality was fair, and it was better than those of the APACHE II score (Fig. 3). The APACHE scoring system is among the most widely used, of which there are four versions (APACHE I through IV scores). Although APACHE IV score is the most up-to-date version, some centres still use older versions including APACHE II score. In the present study, despite having a poor discriminatory ability for predicting hospital and ICU mortalities, an APACHE II score of 21 and above was independently associated with an increased risk of deaths in hospitals (Table 3). However, in contrast to a SOFA score of 10 and above, an APACHE II score of 19 and above was not an independent predictor of ICU mortality (Table 4). Previous studies revealed that the APACHE II score had a good prognostic value in acutely ill or surgical patients (10, 11) but did not differentiate between the sterile and infected necrotizing pancreatitis and had a poor predictive value for the severity of acute pancreatitis at 24 hours (12).

In contrast, the SOFA score was proposed for patients with a suspected infection that an increase of 2 points or more could serve as clinical criteria for sepsis.(1) In ICU patients with suspected infection, discrimination of the SOFA score was fair for predicting hospital mortality, with an AUROC value of 0.74 (95% CI, 0.73 to 0.76; P_AUROC_ <0.001), reported in the previously published studies.(1, 17) However, our study showed that the discriminatory ability of the SOFA score was poor for predicting hospital mortality (Fig. 2). This difference might be due to our SOFA score only calculated upon ICU admission, in contrast to the SOFA score in the previously published study that was calculated for the time window from 48 hours before to 24 hours after the onset of an infection, as well as on each calendar day.(17) This difference also might be because the burden and causes of sepsis and its management differ considerably between HIC and LMIC settings,(7, 35, 37) which might make the accuracy of critical illness severity scoring systems vary widely in the different countries, particularly between HICs and LMICs. However, our study revealed that the SOFA score had a fair discriminatory ability for predicting ICU mortality (Fig. 3). Moreover, a SOFA score of 8 and above and a score of 10 and above were independently associated with an increased risk of deaths in hospitals and ICUs, respectively (Tables 3 and 4). Overall, this study shows that both SOFA and APACHE II scores were worthwhile in predicting hospital and ICU mortalities in ICU patients with sepsis. However, because of having better discrimination for predicting ICU mortality, the SOFA was preferable to the APACHE II score in predicting mortality.

The present study’s data from many centres, which contained few missing data points, was a benefit (Table S19 as shown in Supplementary file 3). The following are some drawbacks of the current study, though: *First*, since there isn’t a national registry of ICUs to enable systematic recruitment of units, we used the snowball method to find suitable units, which may have resulted in the selection of centres with a greater interest in managing sepsis; as a result, our data are subject to selection bias and might not accurately reflect intensive care in all of Vietnam; *Second*, we did not create a protocol for microbiological investigations due to the study’s real-world aspect. The data on point-of-care tests (such as lactate clearance) and life-sustaining therapies (such as fluid balance, steroid administration, and modalities of RRT and MV) were also missing since we primarily evaluated resources used in ICUs. Additionally, we decided not to gather information on antibiotic resistance and appropriateness to increase the practicality of performing the study in busy ICUs; *Thirdly*, the mixed-effects logistic regression model could not be used to predict the discrete outcome variables measured at two different times, i.e., inside and outside the ICU settings, due to our independent variables (e.g., SOFA score that was greater than or equal to the cut-off value), which might be associated with the primary outcome only measured upon ICU admission; *Finally*, even though the sample size was sufficient, the confidence interval was a little bit broad (6.03%), which may have an impact on the sample’s normal distribution. Therefore, more studies with bigger sample sizes may be required to strengthen the findings.

## CONCLUSIONS

Our cohort was a selected population of patients with sepsis admitted to the ICUs in Vietnam with a high mortality rate. The SOFA and APACHE II scores were worthwhile in predicting mortality among ICU patients with sepsis. However, due to better discrimination for predicting ICU mortality, the SOFA score was preferable to the APACHE II score in predicting mortality.

## Data Availability

All data produced in the present study are available upon reasonable request to the authors.

## Data availability statement

Data are available on reasonable request. Data are available on reasonable request to the authors.

## Ethics statements

### Patient consent for publication

Verbal informed consents were directly obtained from patients or, when unavailable, from family members at the ICUs and witnessed by the on-duty medical staff.

### Ethics approval

The Scientific and Ethics Committees of Bach Mai Hospital approved this study (approval number: 2919/QĐ–BM; project code: BM-2017-883-89). We also obtained permission from the heads of institutions and departments of all participating hospitals (see Table S1, as shown in Supplementary file 3, for additional information concerning the hospitals’ names) and their respective institutional review boards wherever available. The study was conducted according to the principles of the Declaration of Helsinki. In this non-intervention study, all collected information has received verbal informed consent from patients or, when unavailable, from family members at the ICUs, and witnessed by the on-duty medical staff. Written informed consent, however, was waived by the Bach Mai Hospital Scientific and Ethics Committees since it was not feasible to undergo such a methodical process of collection when the subject was comprised of an urgent situation in which a patient or a family member’s condition was severe or life-threatening. Public notification of the study was made by public posting. All data analyses were based upon datasets kept in password-protected systems, and all final presented data have been made anonymous.

## Acknowledgments

We thank all ED and ICU staff of participating hospitals for their support with this study. We also thank the staff of the Faculty of Public Health at Thai Binh University of Medicine and Pharmacy for their support and statistical advice. Finally, we thank Miss Phuong Thi Tran from the Center for Emergency Medicine of Bach Mai Hospital, Hanoi, Vietnam, and Miss Mai Phuong Nguyen from St. Paul American School, Hanoi, Vietnam, for their support with this study.

## Supplementary materials

Supplementary file 1.pdf

Supplementary file 2.pdf

Supplementary file 3.pdf

## Authors’ contribution

SND contributed to the conception, design of the work, acquisition, and interpretation of data for the work, and revised the draft critically for important intellectual content; CQL contributed to the conception, design of the work, acquisition, analysis, and interpretation of data for the work, and wrote the first draft of the work; CXD and TAN contributed to the design of the work, acquisition, interpretation of data for the work, and revised the draft critically for important intellectual content; DTP and MHN contributed to the design of the work, analysis, and interpretation of data for the work; NTN, DQH, QTAH, CVB, TDV, HNB, HTN, HBH, TTPL, LTBN, PTD, TDN, VHL, and GTTP contributed to the acquisition and interpretation of data for the work; GTHB, TVB, TTNP, CVN, and ADN contributed to the interpretation of data for the work; JP and AL contributed to the design of the work, interpretation of data for the work, and revised the draft critically for important intellectual content. All authors reviewed and edited the work and approved its final version.

## References

1. Singer M, Deutschman CS, Seymour CW, Shankar-Hari M, Annane D, Bauer M, et al. The Third International Consensus Definitions for Sepsis and Septic Shock (Sepsis-3). Jama. 2016;315(8):801–10.

2. Rudd KE, Johnson SC, Agesa KM, Shackelford KA, Tsoi D, Kievlan DR, et al. Global, regional, and national sepsis incidence and mortality, 1990-2017: analysis for the Global Burden of Disease Study. Lancet. 2020;395(10219):200–11.

3. Liu V, Escobar GJ, Greene JD, Soule J, Whippy A, Angus DC, et al. Hospital deaths in patients with sepsis from 2 independent cohorts. Jama. 2014;312(1):90–2.

4. Sakr Y, Jaschinski U, Wittebole X, Szakmany T, Lipman J, Ñamendys-Silva SA, et al. Sepsis in Intensive Care Unit Patients: Worldwide Data From the Intensive Care over Nations Audit. Open forum infectious diseases. 2018;5(12):ofy313.

5. Torio CM, Moore BJ. National Inpatient Hospital Costs: The Most Expensive Conditions by Payer, 2013: Statistical Brief #204. In: David Knutson, editor. Healthcare Cost and Utilization Project (HCUP) Statistical Briefs. Rockville (MD): Agency for Healthcare Research and Quality (US); 2006.

6. Bauer M, Gerlach H, Vogelmann T, Preissing F, Stiefel J, Adam D. Mortality in sepsis and septic shock in Europe, North America and Australia between 2009 and 2019-results from a systematic review and meta-analysis. Critical care (London, England). 2020;24(1):239.

7. Schultz MJ, Dunser MW, Dondorp AM, Adhikari NK, Iyer S, Kwizera A, et al. Current challenges in the management of sepsis in ICUs in resource-poor settings and suggestions for the future. Intensive care medicine. 2017;43(5):612–24.

8. Levy MM, Fink MP, Marshall JC, Abraham E, Angus D, Cook D, et al. 2001 SCCM/ESICM/ACCP/ATS/SIS International Sepsis Definitions Conference. Intensive care medicine. 2003;29(4):530–8.

9. Bone RC, Balk RA, Cerra FB, Dellinger RP, Fein AM, Knaus WA, et al. Definitions for sepsis and organ failure and guidelines for the use of innovative therapies in sepsis. The ACCP/SCCM Consensus Conference Committee. American College of Chest Physicians/Society of Critical Care Medicine. Chest. 1992;101(6):1644–55.

10. Knaus WA, Draper EA, Wagner DP, Zimmerman JE. APACHE II: a severity of disease classification system. Critical care medicine. 1985;13(10):818–29.

11. Capuzzo M, Valpondi V, Sgarbi A, Bortolazzi S, Pavoni V, Gilli G, et al. Validation of severity scoring systems SAPS II and APACHE II in a single-center population. Intensive care medicine. 2000;26(12):1779–85.

12. Banks PA, Freeman ML. Practice guidelines in acute pancreatitis. The American journal of gastroenterology. 2006;101(10):2379–400.

13. Evans L, Rhodes A, Alhazzani W, Antonelli M, Coopersmith CM, French C, et al. Surviving sepsis campaign: international guidelines for management of sepsis and septic shock 2021. Intensive care medicine. 2021;47(11):1181–247.

14. Vincent JL, Moreno R, Takala J, Willatts S, De Mendonca A, Bruining H, et al. The SOFA (Sepsis-related Organ Failure Assessment) score to describe organ dysfunction/failure. On behalf of the Working Group on Sepsis-Related Problems of the European Society of Intensive Care Medicine. Intensive care medicine. 1996;22(7):707–10.

15. Vincent JL, de Mendonça A, Cantraine F, Moreno R, Takala J, Suter PM, et al. Use of the SOFA score to assess the incidence of organ dysfunction/failure in intensive care units: results of a multicenter, prospective study. Working group on “sepsis-related problems” of the European Society of Intensive Care Medicine. Critical care medicine. 1998;26(11):1793–800.

16. Ferreira FL, Bota DP, Bross A, Mélot C, Vincent JL. Serial evaluation of the SOFA score to predict outcome in critically ill patients. Jama. 2001;286(14):1754–8.

17. Seymour CW, Liu VX, Iwashyna TJ, Brunkhorst FM, Rea TD, Scherag A, et al. Assessment of Clinical Criteria for Sepsis: For the Third International Consensus Definitions for Sepsis and Septic Shock (Sepsis-3). Jama. 2016;315(8):762–74.

18. Shankar-Hari M, Phillips GS, Levy ML, Seymour CW, Liu VX, Deutschman CS, et al. Developing a New Definition and Assessing New Clinical Criteria for Septic Shock: For the Third International Consensus Definitions for Sepsis and Septic Shock (Sepsis-3). Jama. 2016;315(8):775–87.

19. World Bank. World Development Indicators Washington, D.C., United States: The World Bank Group; 2019 [updated February 12, 2021; cited 2022 March 19]. Available from: https://databank.worldbank.org/data/download/POP.pdf.

20. World Health Organization. Viet Nam SARS-free. 2003. In: Weekly epidemiological record [Internet]. Geneva, Switzerland: The World Health Organization; [145-6]. Available from: https://www.who.int/wer/2003/en/wer7818.pdf.

21. South East Asia Infectious Disease Clinical Research Network. Effect of double dose oseltamivir on clinical and virological outcomes in children and adults admitted to hospital with severe influenza: double blind randomised controlled trial. Bmj. 2013;346:f3039.

22. World Health Organization. Cumulative number of confirmed human cases for avian influenza A(H5N1) reported to WHO, 2003-2021, 15 April 2021 Geneva, Switzerland: The World Health Organization; 2021 [updated April 15, 2021; cited 2022 August 31]. Available from: https://www.who.int/publications/m/item/cumulative-number-of-confirmed-human-cases-for-avian-influenza-a(h5n1)-reported-to-who-2003-2021-15-april-2021.

23. World Health Organization. COVID-19 Situation reports in Viet Nam Geneva, Switzerland: The World Health Organization; 2021 [updated December 15, 2021; cited 2021 December 22]. Available from: https://www.who.int/vietnam/emergencies/coronavirus-disease-(covid-19)-in-viet-nam/covid-19-situation-reports-in-viet-nam/.

24. Do TV, Manabe T, Vu GV, Nong VM, Fujikura Y, Phan D, et al. Clinical characteristics and mortality risk among critically ill patients with COVID-19 owing to the B.1.617.2 (Delta) variant in Vietnam: A retrospective observational study. PloS one. 2023;18(1):e0279713.

25. Anders KL, Nguyet NM, Chau NV, Hung NT, Thuy TT, Lien le B, et al. Epidemiological factors associated with dengue shock syndrome and mortality in hospitalized dengue patients in Ho Chi Minh City, Vietnam. The American journal of tropical medicine and hygiene. 2011;84(1):127–34.

26. Mai NT, Hoa NT, Nga TV, Linh le D, Chau TT, Sinh DX, et al. Streptococcus suis meningitis in adults in Vietnam. Clinical infectious diseases : an official publication of the Infectious Diseases Society of America. 2008;46(5):659–67.

27. Maude RJ, Ngo TD, Tran DT, Nguyen BTH, Dang DV, Tran LK, et al. Risk factors for malaria in high incidence areas of Viet Nam: a case-control study. Malaria journal. 2021;20(1):373.

28. Nguyen KV, Thi Do NT, Chandna A, Nguyen TV, Pham CV, Doan PM, et al. Antibiotic use and resistance in emerging economies: a situation analysis for Viet Nam. BMC public health. 2013;13:1158.

29. Phu VD, Wertheim HFL, Larsson M, Nadjm B, Dinh Q-D, Nilsson LE, et al. Burden of Hospital Acquired Infections and Antimicrobial Use in Vietnamese Adult Intensive Care Units. PloS one. 2016;11(1):e0147544.

30. The World Bank. The World Bank In Vietnam. Washington: The World Bank; 2020 [updated October 6, 2020; cited 2022 March 19]. Available from: https://www.worldbank.org/en/country/vietnam/overview.

31. Dat VQ, Long NT, Giang KB, Diep PB, Giang TH, Diaz JV. Healthcare infrastructure capacity to respond to severe acute respiratory infection (SARI) and sepsis in Vietnam: A low-middle income country. Journal of critical care. 2017;42:109–15.

32. Chinh LQ, Manabe T, Son DN, Chi NV, Fujikura Y, Binh NG, et al. Clinical epidemiology and mortality on patients with acute respiratory distress syndrome (ARDS) in Vietnam. PloS one. 2019;14(8):e0221114–e.

33. Takashima K, Wada K, Tra TT, Smith DR. A review of Vietnam’s healthcare reform through the Direction of Healthcare Activities (DOHA). Environmental health and preventive medicine. 2017;22(1):74.

34. Asian Critical Care Clinical Trials Group. Management of Sepsis in Asia’s Intensive Care unitS II (MOSAICS II) Study Singapore: Society of Intensive Care Medicine (Singapore); 2019 [updated October 2019; cited 2021 December 14]. Available from: https://sicm.org.sg/article/YDNc8.

35. Do SN, Luong CQ, Pham DT, Nguyen MH, Nguyen NT, Huynh DQ, et al. Factors relating to mortality in septic patients in Vietnamese intensive care units from a subgroup analysis of MOSAICS II study. Scientific reports. 2021;11(1):18924.

36. Li A, Ling L, Qin H, Arabi YM, Myatra SN, Egi M, et al. Epidemiology, Management, and Outcomes of Sepsis in ICUs among Countries of Differing National Wealth across Asia. American journal of respiratory and critical care medicine. 2022;206(9):1107–16.

37. Do SN, Luong CQ, Nguyen MH, Pham DT, Nguyen NT, Huynh DQ, et al. Predictive validity of the quick Sequential Organ Failure Assessment (qSOFA) score for the mortality in patients with sepsis in Vietnamese intensive care units. PloS one. 2022;17(10):e0275739.

38. Levy MM, Evans LE, Rhodes A. the Surviving Sepsis Campaign Bundle: 2018 update. Intensive care medicine. 2018;44(6):925–8.

39. Thao PTN, Tra TT, Son NT, Wada K. Reduction in the IL-6 level at 24 h after admission to the intensive care unit is a survival predictor for Vietnamese patients with sepsis and septic shock: a prospective study. BMC emergency medicine. 2018;18(1):39.

40. Phua J, Koh Y, Du B, Tang YQ, Divatia JV, Tan CC, et al. Management of severe sepsis in patients admitted to Asian intensive care units: prospective cohort study. Bmj. 2011;342:d3245.

41. Dewi RS, Radji M, Andalusia R. Evaluation of Antibiotic Use Among Sepsis Patients in an Intensive Care Unit: A cross-sectional study at a referral hospital in Indonesia. Sultan Qaboos University medical journal. 2018;18(3):e367–e73.

42. Rudd KE, Hantrakun V, Somayaji R, Booraphun S, Boonsri C, Fitzpatrick AL, et al. Early management of sepsis in medical patients in rural Thailand: a single-center prospective observational study. Journal of Intensive Care. 2019;7(1):55.

43. Rhodes A, Evans LE, Alhazzani W, Levy MM, Antonelli M, Ferrer R, et al. Surviving Sepsis Campaign: International Guidelines for Management of Sepsis and Septic Shock: 2016. Critical care medicine. 2017;45(3):486–552.

44. Phua J, Lim C-M, Faruq MO, Nafees KMK, D. B, Gomersall CD, et al. The story of critical care in Asia: a narrative review. Journal of Intensive Care. 2021;9(1):60.

45. Vincent JL, Sakr Y, Singer M, Martin-Loeches I, Machado FR, Marshall JC, et al. Prevalence and Outcomes of Infection Among Patients in Intensive Care Units in 2017. Jama. 2020;323(15):1478–87.

46. Sharma S, Kelly YP, Palevsky PM, Waikar SS. Intensity of Renal Replacement Therapy and Duration of Mechanical Ventilation: Secondary Analysis of the Acute Renal Failure Trial Network Study. Chest. 2020;158(4):1473–81.

47. Elseviers MM, Lins RL, Van der Niepen P, Hoste E, Malbrain ML, Damas P, et al. Renal replacement therapy is an independent risk factor for mortality in critically ill patients with acute kidney injury. Critical care (London, England). 2010;14(6):R221.

48. Barbar SD, Clere-Jehl R, Bourredjem A, Hernu R, Montini F, Bruyère R, et al. Timing of Renal-Replacement Therapy in Patients with Acute Kidney Injury and Sepsis. New England Journal of Medicine. 2018;379(15):1431–42.

49. Aboshakwa AM, Lalla U, Irusen EM, Koegelenberg CFN. Acinetobacter baumannii infection in a medical intensive care unit: The impact of strict infection control. African journal of thoracic and critical care medicine. 2019;25(1).

50. John AO, Paul H, Vijayakumar S, Anandan S, Sudarsan T, Abraham OC, et al. Mortality from acinetobacter infections as compared to other infections among critically ill patients in South India: A prospective cohort study. Indian journal of medical microbiology. 2020;38(1):24–31.

51. Shargian-Alon L, Gafter-Gvili A, Ben-Zvi H, Wolach O, Yeshurun M, Raanani P, et al. Risk factors for mortality due to Acinetobacter baumannii bacteremia in patients with hematological malignancies -a retrospective study. Leukemia & lymphoma. 2019;60(11):2787–92.

